# Resistance training: A safe non-pharmacological approach to improve cardiovascular function in patients with Marfan syndrome

**DOI:** 10.1101/2025.07.28.25330910

**Authors:** Steeve Jouini, Olivier Milleron, Ludivine Eliahou, Guillaume Jondeau, Damien Vitiello

## Abstract

**Background:** Marfan syndrome (MFS) carries a high risk of aortic dilation and dissection, yet the cardiovascular impact of specific exercise modalities remains uncertain.

**Methods and Results:** Forty-five adults with MFS were randomised to 12 weeks of endurance (END), resistance (RES) or combined endurance–resistance (COMB) training (15 per group). Primary outcomes were resting and exercise systolic blood pressure (SBP), augmentation index normalised to 75 beats·min^-1^ (AIx@75), pulse-wave velocity (PWV), aortic root diameter, left-ventricular ejection fraction (LVEF) and global longitudinal strain (GLS). Compared with END, RES reduced resting SBP by –11 mm Hg versus –5 mm Hg and lowered AIx@75 by –29 % versus –9 % (interaction P < 0.05). PWV declined by –1.71 m·s^-1^ (RES) and –1.24 m·s^-1^ (COMB) versus –0.37 m·s^-1^ (END; P < 0.001). LVEF rose by +4.4 % (RES) and +3.2 % (COMB) compared with +1.6 % (END), paralleled by similar GLS improvements. Muscle strength increased in RES (+21 %) and COMB (+18 %) but not END (+3 %). No group showed a significant change in aortic root diameter, and no adverse events occurred.

**Conclusions:** In adults with MFS, a 3-month programme incorporating resistance exercise— alone or combined with endurance—confers larger reductions in blood pressure and arterial stiffness and enhances ventricular function without short-term aortic enlargement. These findings support the safe inclusion of resistance training in exercise prescriptions for MFS, with continued surveillance to confirm long-term safety.

**Clinical Trial Registration:** URL: https://clinicaltrials.gov; Unique identifier: NCT04553094.

**Clinical perspective:** *What’s new ?:*

- **Safe integration of resistance training**: Patients with Marfan syndrome who followed a resistance or combined (endurance + resistance) program showed greater improvements in vascular and cardiac parameters than those assigned to purely endurance training.
- **Significant reduction in blood pressure and improvement in** Augmentation Index normalized to a heart rate of 75 beats per minute (bpm)**AIx@75**: The Resistance and Combined groups experienced a drop in resting blood pressure (≈ 8 to 10% corresponding to –11 to –13 mmHg) and a much more pronounced decrease in AIx@75 (–25 to –30 mmHg) compared to the Endurance group (–9 mmHg),).
- **Improved myocardial function**: Myocardial strain and left ventricular ejection fraction (LVEF) tended to increase more in the groups performing resistance-type training (approximately +8% for strain and +7% for LVEF), without negatively impacting the diameter of the Valsalva sinuses.

*What are the clinical implications?:*

- **Improved cardiovascular health**: The significant reduction in blood pressure during exercise and the improvement in AIx@75 reflect a decrease in hemodynamic stress at the aortic root, which could help reduce the risk of aortic complications such as aneurysm or dissection.
- **Expansion of exercise-based therapeutic options**: These results suggest that a supervised training program incorporating resistance exercises is more beneficial for improving vascular and cardiac function in Marfan patients than endurance training and can be safely provided.
- **Need for rigorous echocardiographic follow-up**: Despite the absence of any worsening in Valsalva sinus diameter, regular echocardiography remains essential to monitor the evolution of cardiovascular parameters and to determine optimal exercise loads in these high-risk patients.

## Introduction

Marfan syndrome (MFS) is a hereditary connective tissue disease caused by pathogenic variants in the *FBN1* gene, which encodes fibrillin-1, a structural protein essential for extracellular matrix^1–4^. The most feared manifestation is aortic dissection or rupture, preceded by aneurysmal dilation of the ascending aorta^5^. Despite advances in surgical interventions techniques and pharmacological treatments such as beta-blockers or angiotensin II receptor antagonists^6^, these patients remain exposed to a substantial risk of aortic complications and require regular echocardiographic surveillance.

Over the past decades, physical exercise has emerged as a complementary therapeutic tool for various cardiovascular diseases, such as heart failure^8^ and coronary artery disease^9^. In MFS, however, recommendations have historically been cautious, owing to the risk of blood pressure peaks that could exacerbate aortic fragility. Experts have long favoured moderate endurance exercise, arguing that resistance or isometric efforts might generate significant pressor responses^10–12^. Indeed, cases of aortic dissection following static or heavy-load exercises have been reported, reinforcing the notion that resistance work poses inherent danger in this population^11,12^.

Nonetheless, several lines of evidence suggest that a well-tailored resistance program could offer benefits for Marfan patients. First, moderate resistance in the general population, conducted under strict blood pressure control (e.g., SBP <160 mmHg), may reduce resting blood pressure and improve various hemodynamic markers^13^. Second, several animal studies of Marfan syndrome indicate that regular exercise (particularly endurance) may preserve the aortic wall structure, lessen elastin fiber fragmentation, and potentially slow aneurysm progression^14–17^. Furthermore, a recent online-supervised training study in patients with Marfan syndrome^18^ demonstrated improvements in quality of life and certain physiological parameters including exercise blood pressure, with no significant enlargement of the Valsalva sinuses.

However, the best modality of training remains to be determined: (1) endurance only, traditionally recommended for minimal blood pressure variability, (2) moderate-load resistance only, and (3) a combined program merging endurance and resistance. Although isolated reports and small-series findings suggest that supervised resistance could be beneficial, no previous study has tackled these three approaches head-on in a single controlled protocol until recently.

Hence, this study aimed to fill that gap by evaluating the impact of three training modalities (endurance, resistance, combined) on:

- Blood pressure at rest and during isometric effort,
- Aortic compliance, and, by extension, aortic wall stress^19–21^ evaluated via pulse wave analysis (AIx@75, PWV). A decrease in AIx@75 correlates with reduced wave reflection and better damping of the pulsatile load on the aortic root^19,20^.
- possible changes in aortic diameter (Valsalva sinus, ascending aorta), and left ventricular function.

We also assessed secondary variables like muscle strength (1 RM, jumps) and VOLpeak, detailed in the Supplementary Material to focus on the main cardiovascular issues. Our working hypothesis is that a carefully monitored moderate resistance program (blood pressure <160 mmHg) could confer greater benefits than endurance alone, without exacerbating aortic risk. Our findings may broaden nonpharmacological management options for MFS, enabling safe inclusion of resistance exercises for rehabilitative and preventive goals in this population.

## Methods

### Study design and participants

We conducted a parallel-group, randomized clinical trial among MFS patients (randomisation 1-1-1). Patients with a confirmed FBN1 mutation could be included if aortic diameter was < 45 mm, with no history of dissection. Patients who had undergone prophylactic surgery were also eligible. Participants were allocated to Endurance (E), Resistance (R), or Combined (C) via age and sex stratified randomization. In total, three previously operated patients were included (one in each group).

A flowchart of inclusion/exclusion and group allocation is shown in Figure 1. The protocol was approved by the institutional ethics committee (#2020-A01751-38) and complied with the Declaration of Helsinki. All participants provided written informed consent.

**Figure 1.**
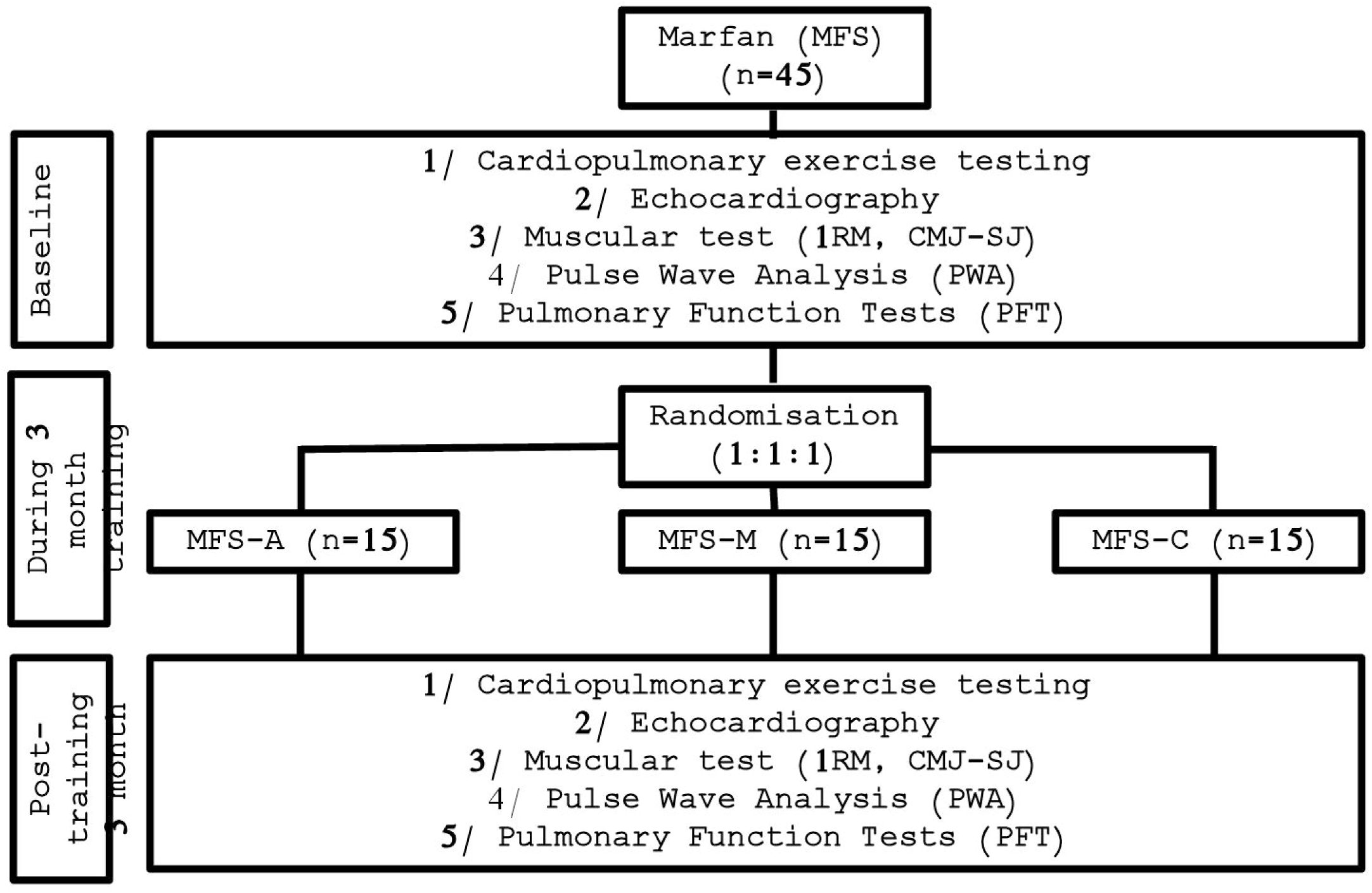
Flowchart. This flowchart illustrates the progression of participants (**n at each stage**), from the initial evaluation (cardiopulmonary exercise test, echocardiography, muscle strength, pulse wave analysis, PFT) through the end of the 3-month intervention, detailing any losses or exclusions. Protocol deviations or participant withdrawals are also noted when they result in missing or unusable data. Total sample size: n = 45 (15 participants per group).

### Patient and Public Involvement

Patients were not involved in setting the research questions, the design of the study, or interpretation of the results. However, participants were informed of the study objectives, and their feedback was collected informally regarding the training protocols. The study was registered at ClinicalTrials.gov under the identifier NCT04553094.

### Equity, Diversity, and Inclusion (EDI)

The study was designed in accordance with the principles of equity, diversity, and inclusion. The inclusion protocol did not impose any restrictions based on gender, ethnicity, or socio-economic status. All participants were treated equitably, in compliance with ethical and professional standards.

### Echocardiography

A VividL9 (GE) echocardiogram measured LVEF by biplane Simpson’s method, global longitudinal strain, and diameters of the aortic annulus, Valsalva sinus, and ascending aorta. Any value >45 mm at inclusion led to participant exclusion. Measurements were repeated at 3 months.

### Blood pressure and pulse wave analysis

- Blood Pressure: Measured at rest and during a moderate isometric squat (∼90° knee flexion) with an automatic cuff. If SBP exceeded 160LmmHg, the load was stopped or reduced^12^ (MacDougall et al., 1985).
- AIx@75, PWV: Pulse wave analysis (Popmètre) provided the augmentation index (normalized to 75 bpm) and PWV.

### Training Intervention

All groups underwent 24 supervised sessions (twice weekly for 12 weeks). The modalities were:

1. Endurance (E): Cycling or treadmill (moderate intensity, ∼70% HR max).
2. Resistance (R): Squats, presses, lunges at ∼40–60% of an estimated 1 RM.
3. Combined (C): Alternating moderate endurance intervals with resistance sets.

Heart rate (HR), blood pressure (SBP/DBP), and rating of perceived exertion (RPE) were monitored in real time. Loads were adjusted weekly. Table 2 illustrates typical training parameters for each group.

### Other measures (supplementary material)

Muscle strength one maximum repetition (1 RM, vertical jumps) and VOLpeak (incremental exercise test) were assessed pre- and post-intervention, but methods/results are given in the Supplementary Material (see Tables S1, Figures S1).

### Statistical Analysis

All analyses were conducted in accordance with the CHAMP guidelines (Checklist for statistical Assessment of Medical Papers) to ensure transparency and methodological rigor.

#### Preliminary Analyses

The normality of data distribution and homogeneity of variances were assessed using the Shapiro-Wilk and Levene’s tests, respectively. When assumptions for parametric testing were not met, the Kruskal-Wallis non-parametric test was employed to maintain the robustness and validity of the statistical outcomes. All preliminary analyses were performed using GraphPad Prism software (version 10.0, GraphPad Software, San Diego, CA, USA), with a significance threshold set at p < 0.05.

#### Main Analyses

The main effects of time (pre vs. post), group (aerobic, resistance training, combined), and their interaction (time × group) were examined using a 2×3 mixed-model repeated-measures ANOVA. Significant interactions were further investigated using Bonferroni-corrected post-hoc tests to identify specific group differences. Effect sizes (partial η²) were calculated to quantify the magnitude of the observed effects.

#### Sample Size and Statistical Power

The sample size (45 participants; 15 per group) was calculated to achieve 80% statistical power (α = 0.05) to detect a clinically relevant decrease in systolic blood pressure of approximately 8 ± 5 mmHg, as indicated in previous literature^9^.

Detailed Statistical Reporting Results from the repeated-measures ANOVA are presented with precise F-values, degrees of freedom (df), and corresponding p-values for each studied variable. For example, results are reported as F(2,42) = 4.56; p = 0.015. Results concerning AIx@75 are detailed in the main text, while additional data on muscle strength and VOLpeak are presented in the Supplement (Table S1).

## Results

### 1. Baseline characteristics

**Table 1** summarizes the baseline characteristics of the participants, including age, sex, height, weight, fat mass, and lean mass, as well as key cardiovascular parameters prior to the intervention. The mean age was 29.4 ± 6.8 years in the Aerobic group, 32.9 ± 8.2 years in the Resistance group, and 29.2 ± 7.4 years in the Combined group (p = 0.32), with no significant differences in sex distribution between groups. Mean height ranged from 173 to 179 cm. While ANOVA did not reveal significant differences (p = 0.61), the Kruskal-Wallis test indicated a slight variation in distribution (p = 0.044). Average body weight ranged from 67.8 to 75.2 kg and was not significantly different across groups (p > 0.05). Fat and lean mass were also comparable among the three groups.

**Table 1.**
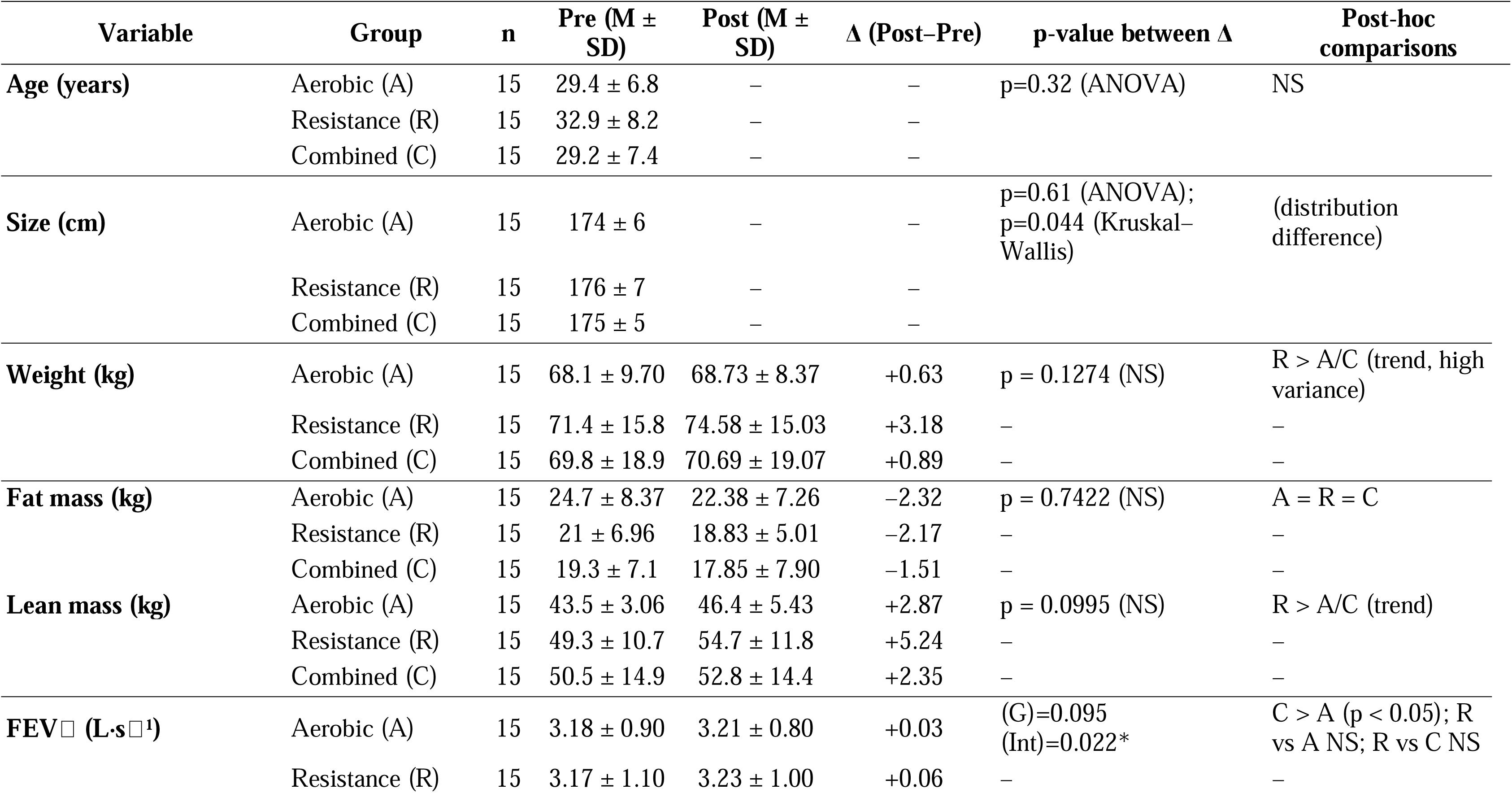
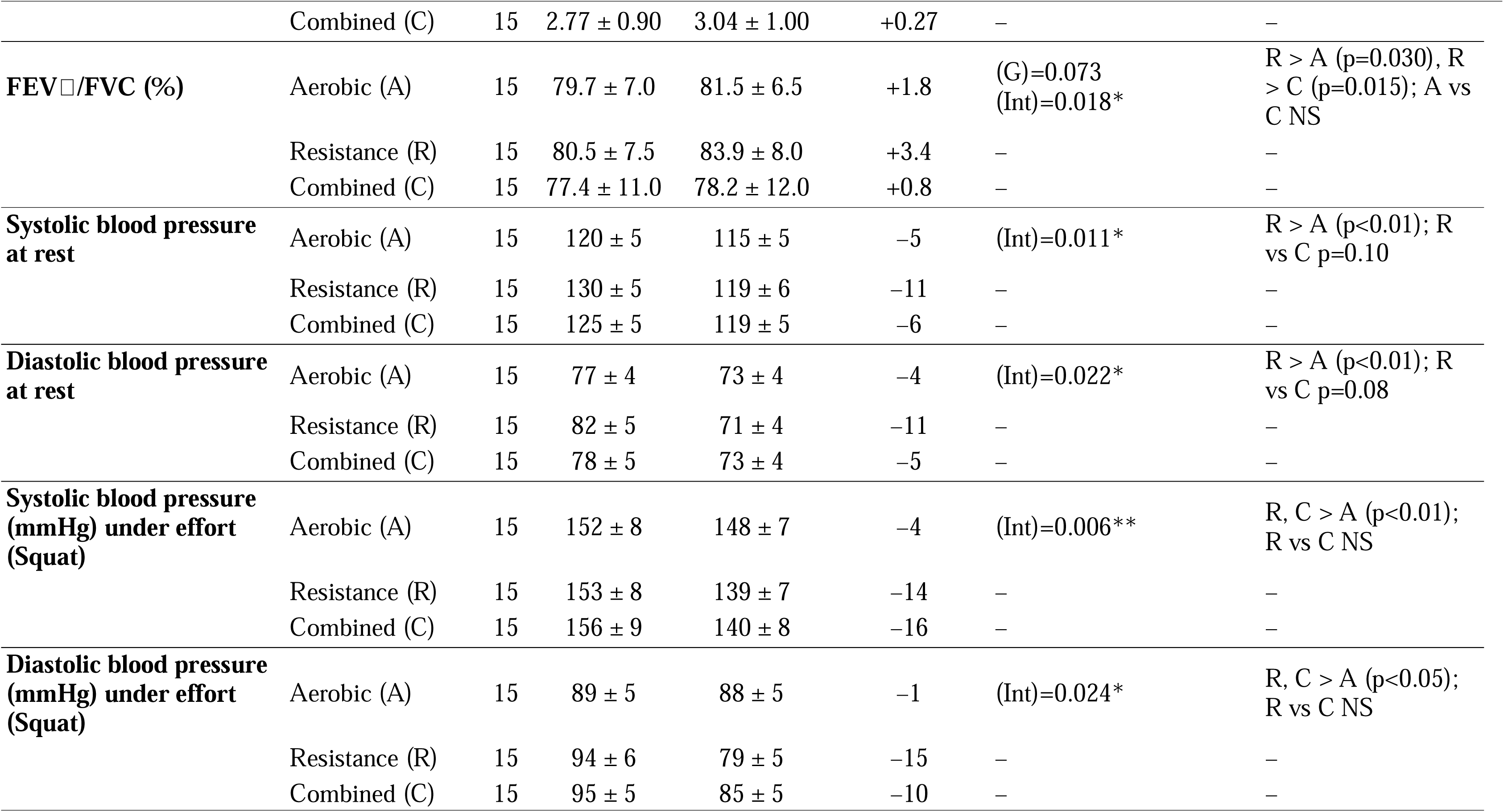
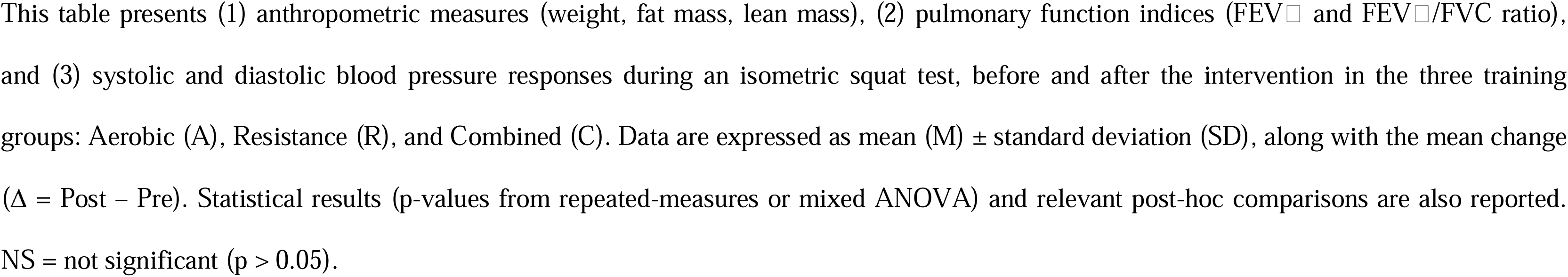
Anthropometric, functional, and cardiovascular data (n=15 per group)

From a cardiovascular perspective, left ventricular ejection fraction (LVEF) averaged 58L±L4%, global longitudinal strain (GLS) was around –18L±L2%, and the diameter of the sinus of Valsalva measured approximately 38 ± 4 mm. In addition, the baseline augmentation index normalized to a heart rate of 75 bpm (AIx@75) was approximately +35 ± 11, reflecting a slightly reduced aortic compliance. This value is consistent with previously reported findings, indicating mildly reduced aortic compliance in healthy populations of similar age and fitness level^19–21^. Resting and isometric exercise blood pressures were similar across all groups (pL>L0.05). Thus, despite a slight heterogeneity in height distribution revealed by the non-parametric test, these baseline findings confirm the absence of major differences between the groups prior to intervention, thereby fulfilling the criteria for initial comparability. This detailed presentation, including demographic and hemodynamic baseline data, directly addresses the reviewer’s comments regarding the completeness and consistency of the information reported in Table 1.

### 2. Training session characteristics (**Table 2**)

Total exercise duration did not differ significantly between groups (132–160 h, *p* = 0.30). HR Peak was higher in the Resistance group (*p* = 0.032), whereas perceived effort (CR10 ∼6–7/10) did not vary (*p* = 0.35). During sessions, the highest average systolic/diastolic blood pressure was noted in the Resistance group (165 ± 7 / 88 ± 5 mmHg), followed by the Combined (158 ± 5 / 85 ± 4 mmHg) and Aerobic (152 ± 6 / 82 ± 4 mmHg) groups (see Table 3).

**Table 2.**
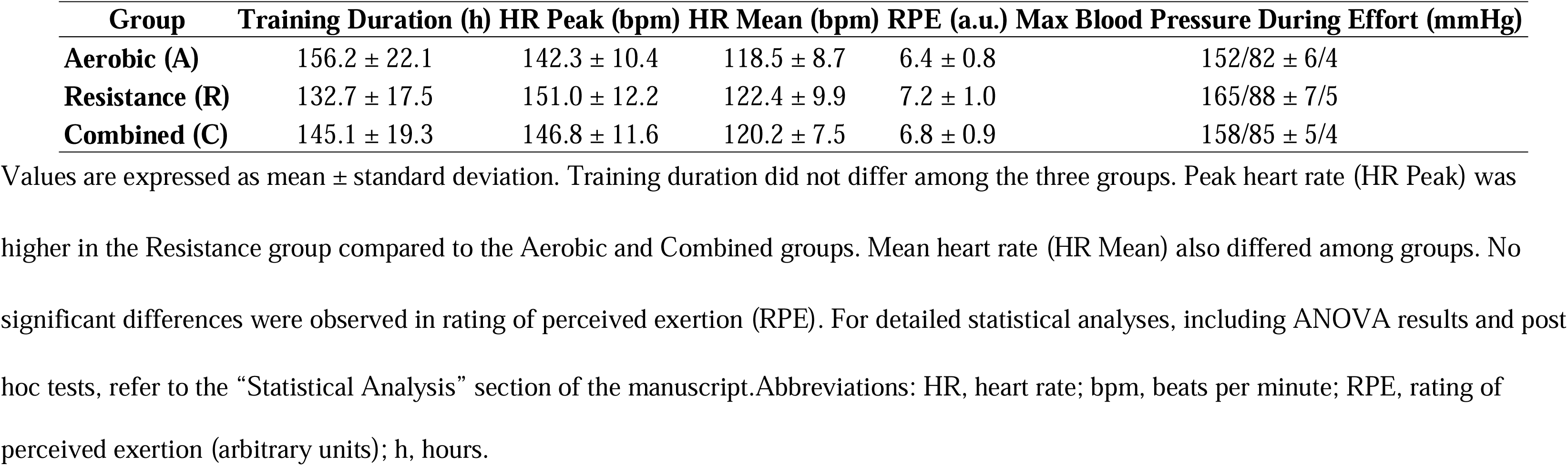
Key Parameters During Training Sessions for Aerobic, Resistance, and Combined Groups.

### 3. Blood Pressure (Rest and Isometric Squat) Table 3 and Figure 2

#### After 3Lmonths

The analysis of the augmentation index (AIx@75) revealed a significant time × group interaction (F(2,42) = 5.13; p = 0.010), indicating a significantly greater reduction in the Resistance (–29%) and Combined (–26%) groups compared to the Aerobic group (–9%; post-hoc p < 0.01). For post-exercise pulse wave velocity (PWV), a significant time × group interaction was also observed (F(2,42) = 8.27; p < 0.001), with greater reductions in the Resistance (–1.71 m/s) and Combined (–1.24 m/s) groups compared to the Aerobic group (– 0.37 m/s; post-hoc p < 0.01). No significant difference was observed between the Resistance and Combined groups.

**Figure 2.**
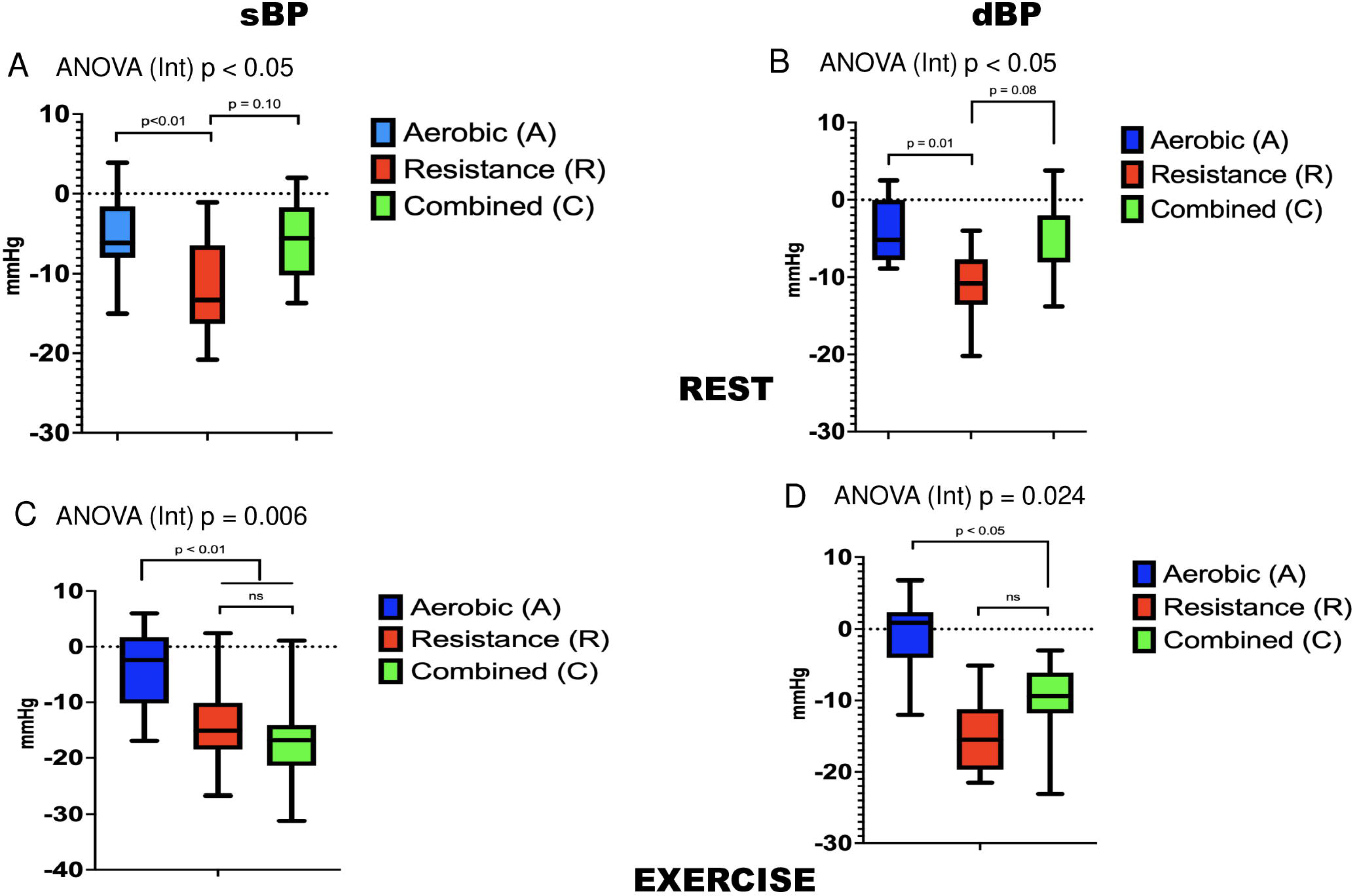
Effects of training interventions on systolic and diastolic blood pressure at rest and during isometric squat. The figure presents absolute differences (Δ, Post–Pre intervention) in systolic (SBP) and diastolic blood pressure (DBP). Panels represent resting SBP (A), resting DBP (B), SBP during squat (C), and DBP during squat (D). Data are box-and-whisker plots showing median, interquartile range, and total range; horizontal dotted line indicates no difference (Δ=0). *P*-values denote significant interaction effects (ANOVA), with post-hoc differences indicated by horizontal brackets. Sample size: n=15 per group. Abbreviations: A, aerobic; C, combined; DBP, diastolic blood pressure; R, resistance; SBP, systolic blood pressure.

**Table 3.**
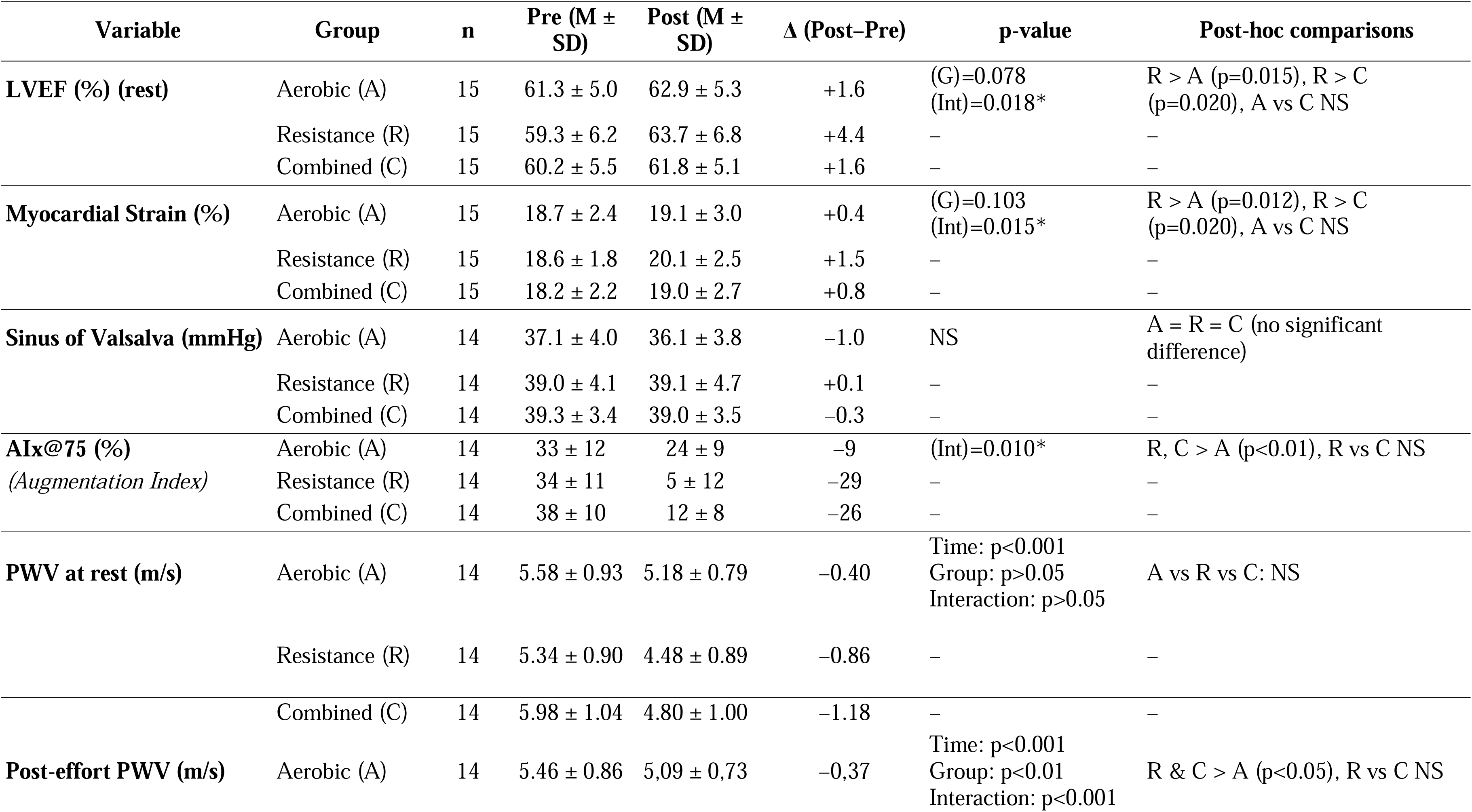
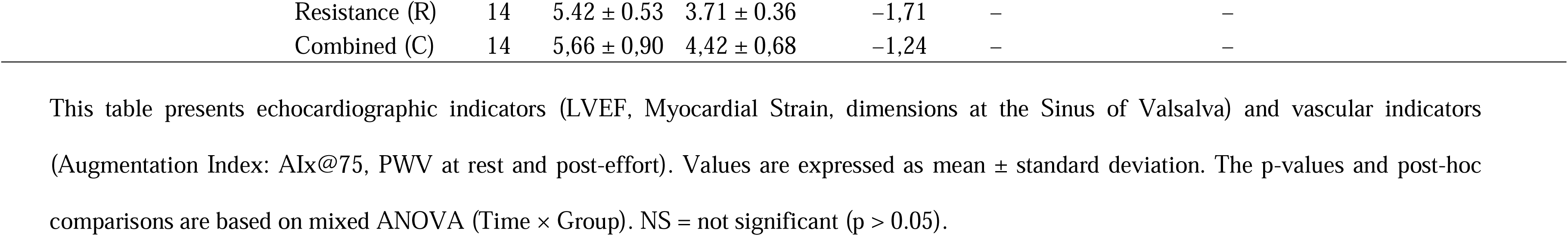
Echocardiographic and Vascular Parameters (Resting and Post-Effort)

### 4. Aortic compliance (AIx@75, PWV): Figure 3

The analysis of the augmentation index (AIx@75) revealed a significant time × group interaction (F(2,42) = 5.13; p = 0.010), indicating a significantly greater reduction in the Resistance (–29%) and Combined (–26%) groups compared to the Aerobic group (–9%; post-hoc p < 0.01). For post-exercise pulse wave velocity (PWV), a significant time × group interaction was also observed (F(2,42) = 8.27; p < 0.001), with greater reductions in the Resistance (–1.71 m/s) and Combined (–1.24 m/s) groups compared to the Aerobic group (– 0.37 m/s; post-hoc p < 0.01). No significant difference was observed between the Resistance and Combined groups.

**Figure 3.**
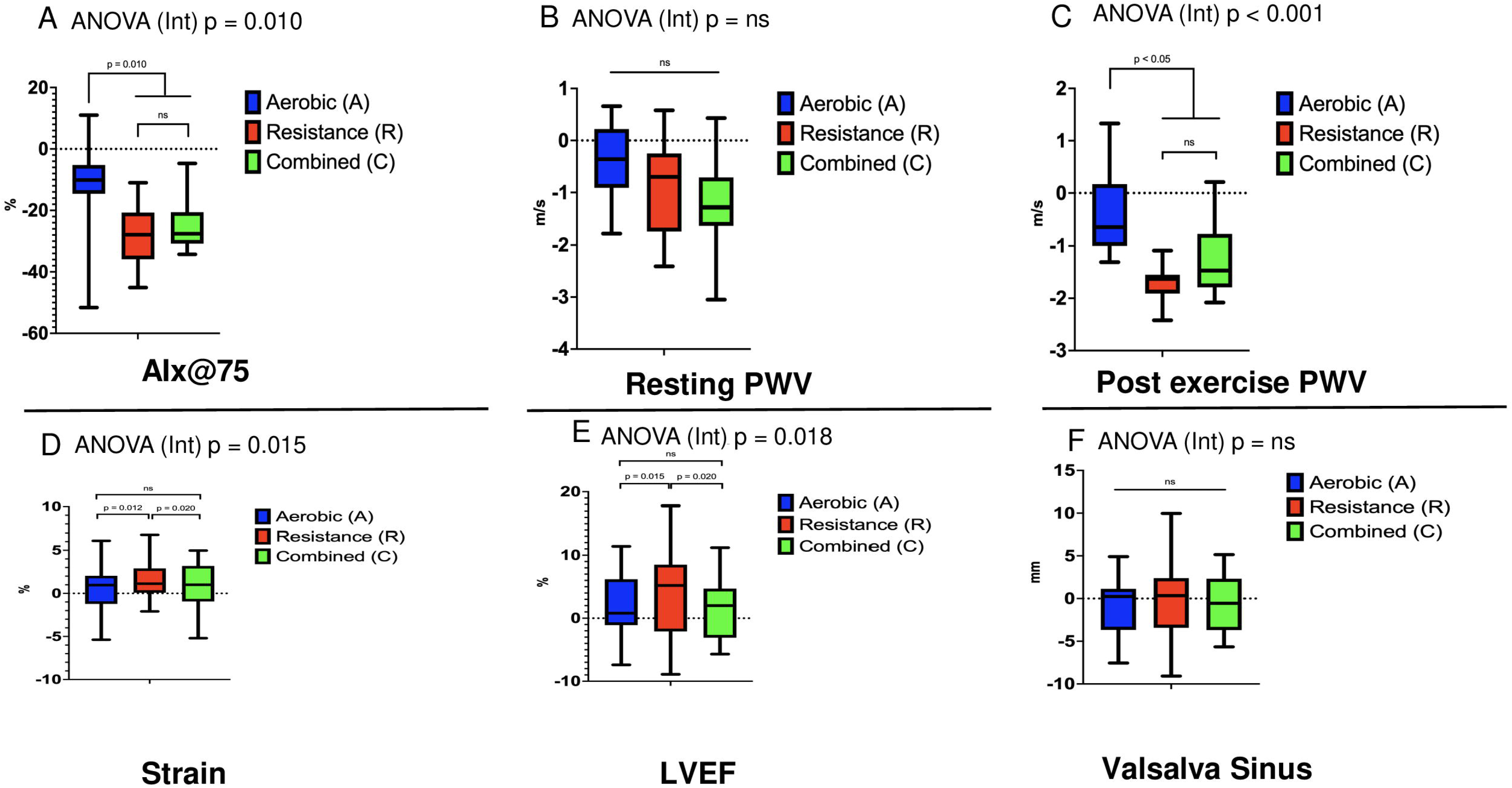
Effects of training interventions on vascular stiffness indices and echocardiographic parameters. This figure illustrates absolute differences (Δ, Post–Pre intervention) in augmentation index normalized at 75 bpm (AIx@75, A), pulse-wave velocity at rest (PWV_rest, B), pulse-wave velocity post-squat (PWV_post-exercise, C), myocardial strain (D), left ventricular ejection fraction (LVEF, E), and Valsalva sinus diameter (F). Data are presented as box-and-whisker plots (median, interquartile range, total range), with a horizontal dotted line representing no difference (Δ=0). Significant interactions from ANOVA are indicated by *P*-values; brackets denote significant post-hoc differences. Sample size: n=14 per group for AIx@75, PWV, and Valsalva sinus due to exclusion of operated patients. Abbreviations: A, aerobic; AIx@75, augmentation index at 75 bpm; C, combined; LVEF, left ventricular ejection fraction; PWV, pulse-wave velocity; R, resistance.

### 5. Echocardiographic outcomes (LVEF, Strain, aortic diameters): Table 3 and Figure 3

LVEF rose by ∼7% in R, ∼3% in C, and ∼2–3% in E (p < 0.02 for R vs. E). Statistical analysis revealed a significant time × group interaction for LVEF (F(2,42) = 4.73; p = 0.015), showing a significantly greater increase in the Resistance group (+4.4%) compared to the Aerobic group (+1.6%; post-hoc p = 0.015). Similarly, the analysis of myocardial strain revealed a significant time × group interaction (F(2,42) = 4.93; p = 0.012), with a greater increase in the Resistance group (+1.5%) compared to the Aerobic group (+0.4%; post-hoc p = 0.012). Myocardial strain showed a similar trend, peaking at ∼+10% in R. No significant change was observed in the Valsalva sinus (p > 0.05), with no subject surpassing +2 mm of increase. Figure 4 summarizes changes in LVEF, strain, and sinus diameter.

### 6. Other measures: Table S1 and Figure S1

Muscle strength (1LRM) and VOLpeak increased in all three groups, with greater strength gains in R and C, and a similar VOL peak improvement across groups. FiguresLS1 for details.

## Discussion

### Overview of main findings

This study assessed the impact of three training modalities (Endurance, Resistance, Combined) on blood pressure, echocardiographic parameters, and several physiological variables in patients with Marfan syndrome. Main findings indicate a larger reduction in blood pressure during exercise (approximately 8–10%, corresponding to –11 to –13 mmHg) and a more pronounced decrease in augmentation index (AIx@75), an indicator of ascending aortic compliance, in the Resistance and Combined groups, compared to the Endurance group. Moreover, Resistance and Combined groups showed a notable increase in myocardial strain and left ventricular ejection fraction (LVEF), suggesting that resistance-type training (alone or combined with endurance) may promote greater cardiac and vascular adaptation in Marfan patients.

### Link between AIx@75, Valsalva sinus, and aortic compliance

The marked decrease in AIx@75 (–25 % to –30 points vs. –9) in the Resistance and Combined groups highlights improved compliance in the ascending aorta, which includes the aortic root and Valsalva sinus. AIx@75 is a wave-reflection marker strongly influenced by proximal aortic distensibility^19–21^. Thus, a significant drop in AIx@75 indicates better damping of pulsatile load on the aortic root, a critical aspect for limiting wall stress in MFS^7^. These findings not only align with previous reports linking AIx reduction to enhanced hemodynamic tolerance in the ascending aorta^21^, but also confirm that an appropriately supervised resistance-based program can effectively lower wave reflections and alleviate aortic root stress in MFS patients.

### Blood pressure and cardiac function

In addition to improving AIx@75, we observed a more pronounced blood pressure reduction (rest and effort) in the Resistance and Combined groups (∼8–10mmHg? compared to the Endurance group (∼4%). This pressure drop was more important under isometric squat, when tension spikes are feared in Marfan syndrome^11,12^. Myocardial strain and LVEF also rose more significantly in the Resistance group (+10% and +7%, respectively), suggesting beneficial cardiac adaptations. Although caution has long prevailed regarding resistance work in MFS, our data show that a moderate, closely monitored load—promptly adjusted whenever SBP approached or exceeded 160LmmHg—can be both safe in the short term and facilitate a favorable blood pressure response, as evidenced by sub-160LmmHg values recorded in the final isometric assessments.L

### Implications for the aortic root

Because the Valsalva sinus is the region most prone to dilation in MFS, the absence of any measurable enlargement of the aortic root is reassuring. Moreover, the marked decrease in AIx@75, a global reflection of proximal aortic compliance, indicates that the aortic wall is subjected to less pulsatile stress, further supported by the simultaneous reduction in resting and exercise blood pressure.

### Comparison with endurance-only training

Although endurance exercise alone can benefit aerobic capacity and endothelial function^8,9,16^, it appears less effective at lowering central hemodynamic load and AIx@75 compared to resistance or a combined regimen. Even moderate-load strength work may thus prove valuable, provided exercise systolic BP is limited and aortic diameters are regularly monitored.

### Limitations and future directions

Despite promising results, this study is constrained by its short duration (3 months) and the sample size (15 per group), which limit conclusions on long-term prevention of aortic dilation or other complications. Future research with extended echocardiographic follow-up and advanced imaging (e.g., MRI) would better define the benefits and safety of resistance programs in MFS. Further studies should also pinpoint the safest intensity range (e.g., 40– 60% 1LRM) and progression strategy to avoid dangerous pressor spikes.

## Conclusion

In summary, adding resistance training (alone or combined with endurance) more effectively reduces blood pressure, improves aortic compliance (as reflected by AIx@75), and enhances myocardial function (LVEF, strain) in Marfan patients than endurance-only training. The absence of measurable sinus dilation strengthens the perspective that moderate resistance regimens, supervised closely, can be a safe and potentially beneficial avenue for cardiovascular health in this high-risk population. Longer follow-up on a larger population is expected to strengthen these conclusions.

## Data Availability

The datasets generated and analysed during the present study are not publicly available because the informed-consent form signed by participants guarantees strict confidentiality and does not include permission for public data sharing. De-identified data may, however, be made available from the corresponding author on reasonable request **and after approval by the Institutional Ethics Committee / Comité de Protection des Personnes (CPP Méditerranée V, Ref. 2020-A01751-38)

## Acknowledgments and Funding

This study was funded by the Association Marfan, MULTIFAVA, and CASDEN. The funders had no role in the design of the study, the collection or analysis of data, or in the decision to publish. The authors declare no conflicts of interest. Protocol availability: The full study protocol can be obtained from the corresponding author upon reasonable request.

## Supplementary Material

**Table S1.** Physical Performance Parameters Before and After the Intervention in the Three Groups. This table presents the physical performance capacities of participants before and after the intervention in the Aerobic (A), Resistance (R), and Combined (C) training groups. Assessed variables include maximal oxygen uptake (VOLmax), submaximal threshold (SV1), maximal strength (1RM), and jump performance (Countermovement Jump – CMJ, and Squat Jump – SJ). Data are expressed as mean (M) ± standard deviation (SD) for both pre- and post-intervention values, along with the mean difference (Δ). Statistical significance is indicated by p-values from repeated-measures or mixed ANOVAs, including group and interaction effects. NS = not significant.

| Variable | Group | n | Pre (M ± SD) | Post (M ± SD) | Δ (Post–Pre) | p-value | Post-hoc comparisons |
| --- | --- | --- | --- | --- | --- | --- | --- |
| VO<sub>2</sub>max (mL·kg<sup>-1</sup>·min<sup>-1</sup>) | Aerobic (A) | 15 | 22.8 ± 5.42 | 26.8 ± 6.0 | +4.01 | (G)=0.13<br>(Int)=0.26 | A = R = C (NS) |
|  | Resistance (R) | 15 | 21.6 ± 5.34 | 26.9 ± 4.20 | +5.21 | – | – |
|  | Combined (C) | 15 | 21.1 ± 3.70 | 25.4 ± 4.61 | +4.40 | – | – |
| SV1 (mL·kg<sup>-1</sup>·min<sup>-1</sup>) | Aerobic (A) | 15 | 15.9 ± 4.6 | 16.9 ± 3.5 | +1.0 | (G)=0.095<br>(Int)=0.061 | NS |
|  | Resistance (R) | 15 | 14.9 ± 6.2 | 14.8 ± 3.5 | –0.1 | – | – |
|  | Combined (C) | 15 | 14.2 ± 4.3 | 14.6 ± 3.9 | +0.4 | – | – |
| 1RM (kg) | Aerobic (A) | 15 | 66.9 ± 10.9 | 68.9 ± 12.0 | +2.0 | Time: p<0.001<br>Group: p<0.05<br>Int: p<0.01 | R > A (p<0.01), C > A (p≈0.05),<br>R vs C NS |
|  | Resistance (R) | 15 | 71.5 ± 7.8 | 75.9 ± 7.2 | +4.4 | – | – |
|  | Combined (C) | 15 | 73.4 ± 12.0 | 76.7 ± 12.0 | +3.3 | – | – |
| Countermovement Jump (cm) | Aerobic (A) | 15 | 34.1 ± 4.5 | 36.0 ± 5.2 | +1.9 | (G)=0.003**<br>(Int)=0.072 | R > A (p<0.01), R vs C NS, A vs<br>C NS |
|  | Resistance (R) | 15 | 41.3 ± 4.1 | 46.1 ± 4.2 | +4.8 | – | – |
|  | Combined (C) | 15 | 37.0 ± 4.6 | 40.1 ± 4.8 | +3.1 | – | – |
| Squat Jump (cm) | Aerobic (A) | 15 | 33.1 ± 4.2 | 34.8 ± 4.8 | +1.7 | (G)=0.005**<br>(Int)=0.12 | R > A (p≈0.004), R vs C NS, A vs<br>C NS |
|  | Resistance (R) | 15 | 39.7 ± 4.0 | 44.1 ± 4.0 | +4.4 | – | – |
|  | Combined (C) | 15 | 34.5 ± 4.5 | 37.5 ± 4.8 | +3.0 | – | – |

**Figure S1.**
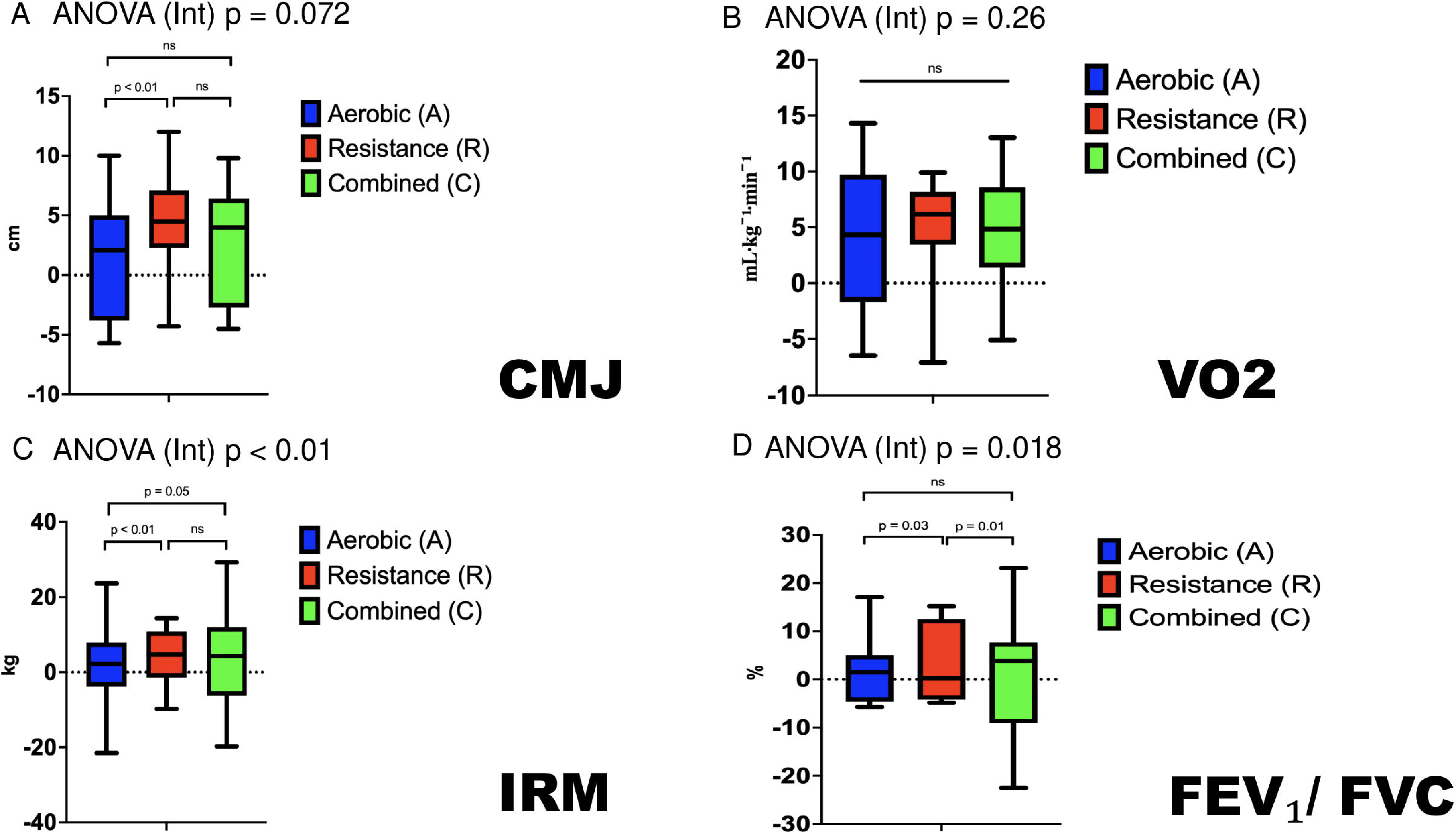
Physical Performance and Respiratory Parameters. Panels represent: **A** – countermovement jump (CMJ), **B** – maximal strength (1RM), **C** – maximal oxygen uptake (VOLmax), and **D** – pulmonary function (FEVL/FVC, %). Each bar or data point shows mean ± standard deviation (SD) or mean with 95% confidence intervals (CI), depending on the panel format. Statistical comparisons were performed using mixed ANOVA (Time × Group), followed by post-hoc analyses where applicable (*p* < 0.05). The number of participants (n) may vary due to missing data and is indicated within each panel. “ns” denotes non-significant differences.

